# A spatiotemporal model of multi-marker antimalarial resistance

**DOI:** 10.1101/2023.10.03.23296508

**Authors:** Yong See Foo, Jennifer A. Flegg

## Abstract

The emergence and spread of drug-resistant *Plasmodium falciparum* parasites has hindered efforts to eliminate malaria. Monitoring the spread of drug resistance is vital, as drug resistance can lead to widespread treatment failure. We develop a Bayesian model to produce spatiotemporal maps that depict the spread of drug resistance, and apply our methods for the antimalarial sulfadoxine-pyrimethamine. We infer from genetic count data the prevalences over space and time of various malaria parasite haplotypes associated with drug resistance. Previous work has focused on inferring the prevalence of individual molecular markers. In reality, combinations of mutations at multiple markers confer varying degrees of drug resistance to the parasite, indicating that multiple markers should be modelled together. However, the reporting of genetic count data is often inconsistent as some studies report haplotype counts, whereas some studies report mutation counts of individual markers separately. In response, we introduce a latent multinomial Gaussian process model to handle partially-reported spatiotemporal count data. As drug-resistant mutations are often used as a proxy for treatment efficacy, point estimates from our spatiotemporal maps can help inform antimalarial drug policies, whereas the uncertainties from our maps can help with optimising sampling strategies for future monitoring of drug resistance.

## 1 Introduction

Malaria is a deadly disease caused by parasites that are transmitted by mosquitoes. During the treatment of a malaria infection, the parasites undergo selective pressure, favouring the survival of parasites that have genetic mutations which confer them resistance against the drug treatment. For *Plasmodium falciparum*, the most common species of malaria parasites, drug resistance is a major threat to the control of the disease. It is therefore important to be able to quantify changes in antimalarial drug resistance, including for sulfadoxine-pyrimethamine (SP) that is used for intermittent preventive treatment for high-risk groups, namely pregnant people and young children.

Spatiotemporal trends in antimalarial drug resistance can be monitored using molecular markers known to be associated with drug resistance as a proxy of clinical efficacy. *P. falciparum* resistance against SP is characterised by mutations on the *dhps* and *dhfr* genes [1]. In fact, SP-resistant parasites often carry multiple SP-resistant mutations (a set of molecular markers or a haplotype). Genetic studies are easier to conduct and are a fraction of the cost of a clinical study — allowing for larger numbers of samples to be collected across more spatiotemporal locations [2]. This makes data from genetic studies readily amenable to model-based geostatistics.

To our knowledge, all works to date that have statistically mapped the geospatial distribution of the *dhps* and *dhfr* markers have modelled each marker separately [3–6]. Most relevant to the work presented in our paper, Flegg et al. [3] developed a predictive model for the geographical and temporal trends across Africa of the prevalence of mutations on the *dhps* gene of the parasite that are associated with SP resistance. A separate model was used for each marker (*dhps* A437G, *dhps* K540E and *dhps* A581G), which models the count data with binomial distributions. Correlation between binomial probabilities are specified according to the spatiotemporal distance between their corresponding sites. Specifically, the logit transformation of the binomial probabilities are set to follow a Gaussian process (GP) distribution.

Although existing spatiotemporal mapping work has focused on individual marker mutations, molecular studies have shown that it is the presence of the double mutant haplotype *dhps* A437G/K540E and triple mutant haplotype *dhps* A437G/K540E/A581G in the parasite that are most strongly associated with an increased risk of SP treatment failure [7, 8], and thus the most clinically relevant. Therefore, new modelling approaches are needed to obtain spatially continuous maps of haplotype prevalences, not just of individual marker mutation prevalences. However, not all studies report the presence or absence of each mutation simultaneously, i.e. the counts of full haplotypes are not reported. Instead, studies may only report the number of samples that carry each individual marker mutation. Since the samples corresponding to each reported count may overlap, we cannot use a multinomial distribution directly. Moreover, some studies only report on a smaller subset of all mutations of interest. We handle these discrepancies caused by partially-reported data under a *latent multinomial model* [9, 10], where the observed counts are treated as binary combinations of unobserved multinomial counts. In this paper, we extend the spatiotemporal GP models of individual *dhps* markers [3, 6] to model the prevalences of multiple haplotypes by using a latent multinomial distribution with GPs. Handling all haplotypes within one model allows us to leverage all available data to greater utility.

This paper is structured as follows. In Section 2, we present the latent multinomial GP model for mapping SP drug resistance based on mutations on the *dhps* gene. We do not account for mutations on the *dhfr* gene as mutations on the *dhps* gene are more closely related to clinical SP failure and there is a triple *dhfr* mutation widely spread across Africa already [11]. This is followed by the outputs on the model in Section 3, showing the prevalence of each haplotype of interest over space and time. Finally, in Section 4 we discuss the implications of our findings.

## 2 Methods

In this paper, we construct a Bayesian hierarchical model that is capable of modelling partially-reported multinomial count data for spatiotemporal changes in drug resistance. This modelling framework is needed to handle reporting inconsistencies found across studies on the prevalences of drug-resistant haplotypes, where different studies may report on different combinations of mutations. For example, the list of *dhps* mutations compiled by the Worldwide Antimalarial Resistance Network [12] includes: *dhps* 437G, *dhps* 540E, *dhps* 581G, *dhps* 437G-540E, *dhps* 437G-540E-A581, and *dhps* 437G-540E-581G. These inconsistencies in data collection and study design significantly complicate model construction and parameter inference. We develop a latent multinomial model (Section 2.1) with a GP prior specification (Section 2.2). We describe how spatiotemporal maps of haplotype prevalences are produced under a Bayesian framework in Section 2.3.

### 2.1 Latent multinomial model formulation

Suppose that we have *G* molecular markers of interest for monitoring drug-resistance. For our application, we have *G* = 3 molecular markers, namely *dhps* 437, *dhps* 540, and *dhps* 581. Define a *full haplotype* to be a binary string of length *G* recording whether each of these mutations are present (1) or absent (0). This results in *H* = 2^*G*^ possible full haplotypes.

Over space and time we have *N* studies (see Figure 1); study *i* (*i* = 1, …, *N*) is conducted to estimate the prevalence **p**_*i*_ at this location of the *H* haplotypes associated with drug resistance. We define **z**_*i*_ to be a vector of length *H* that stores the number of each of the *H* full haplotypes for the *i*th study, and *n*_*i*_ = *z*_*i*,1_ + *z*_*i*,2_ + … + *z*_*i,H*_ to be the known sample size of the study. The counts 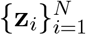 are considered unobserved (latent), each following a multinomial distribution

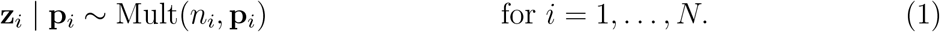

**Figure 1.**
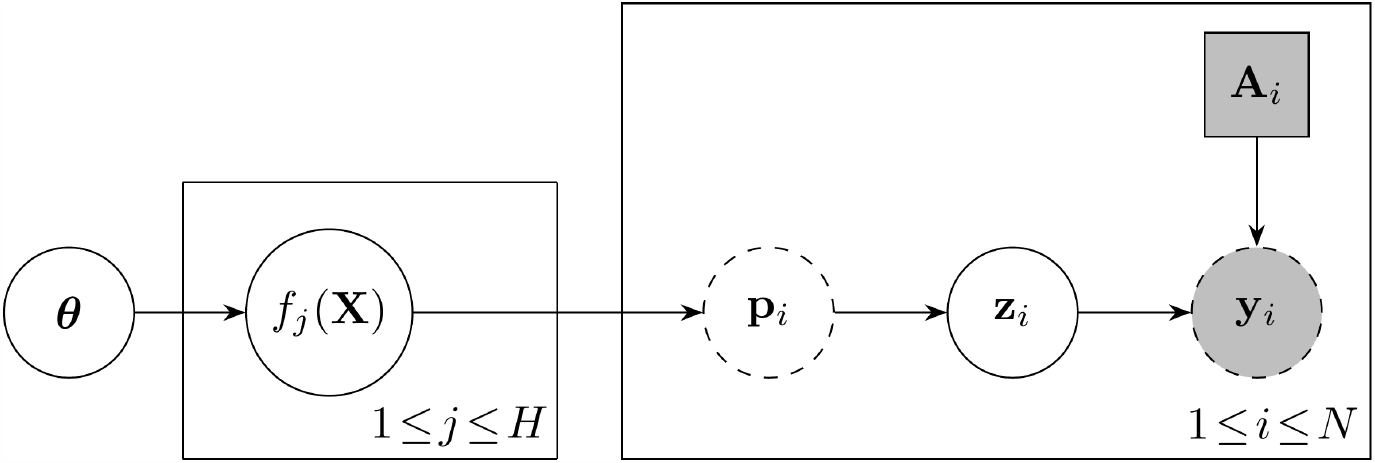
Graphical model for latent multinomial data with multiple populations whose haplo-type prevalences 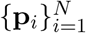 are correlated through Gaussian processes. *f*_*j*_(**X**) denotes the vector (*f*_*j*_(**x**_1_), …, *f*_*j*_(**x**_*N*_)). Circles correspond to random variables while squares correspond to constant values. A shaded node indicates that the variable is observed. A dotted outline indicates that the variable is deterministically calculated from its parent variables. Variables contained within a plate are replicated according to the index at the bottom right.

For each *i* = 1, …, *N*, we observe binary combinations of the latent counts, depending on the way counts are reported in the *i*th study. We refer to the set of full haplotypes that are included by a single observed count as a *realised haplotype*. For example, the realised haplotype 437G-540E includes the full haplotypes 110 (437G-540E-A581) and 111 (437G-540E-581G). The collection of realised haplotypes reported may vary across studies, which we encode within a *configuration matrix*. As an example, for a study which reports on the realised haplotypes *dhps* 437G, *dhps* 540E, *dhps* 581G, *dhps* 437G-540E, *dhps* 437G-540E-A581, and *dhps* 437G-540E-581G, we have the following configuration matrix:

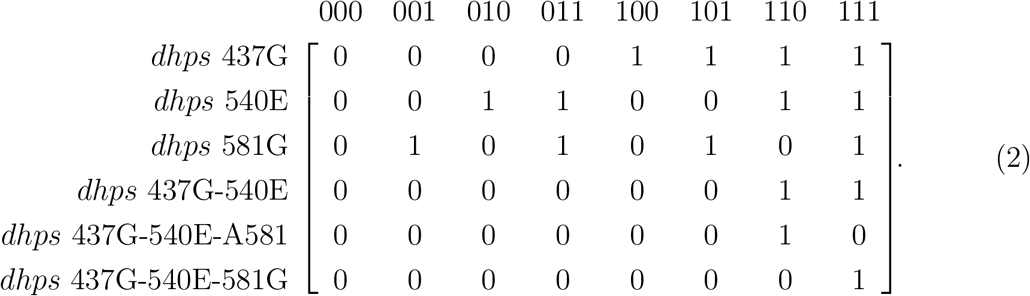

In this way, for an observed count of the samples with the *dhps* 437G mutation, we need to sum the number of the four haplotypes *{*100, 101, 110, 111*}*, while for an observed count of the samples with dhps 437G-540E we only need to sum the haplotypes *{*110, 111*}*. The configuration matrix is constructed based on which haplotypes are included in each count reported by a study.

We denote the number of realised haplotypes reported in study *i* by *R*_*i*_ and the reported counts of the realised haplotypes by vector **y**_*i*_ of length *R*_*i*_, for *i* = 1, …, *N*. Let **A**_*i*_ be the constructed *R*_*i*_ *× H* configuration matrix of 0s and 1s, determined from the realised haplotypes reported in study *i*. For each *i* = 1, …, *N* (see Figure 1), the latent counts **z**_*i*_ must be a nonnegative integer solution to the system

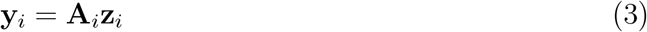

and the sample size constraint *n*_*i*_ = *z*_*i*,1_ + *z*_*i*,2_ + … + *z*_*i,H*_. Since the sample size, *n*_*i*_, is known, we let the first row of **A**_*i*_ always be a vector of ones and the first observed count be the sample size, i.e. *y*_*i*,1_ = *n*_*i*_ for all *i*.

To calculate the likelihood, we marginalise out the latent counts 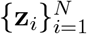 exactly by enumerating all possible latent counts **z**_*i*_ that satisfy (3). The full likelihood is

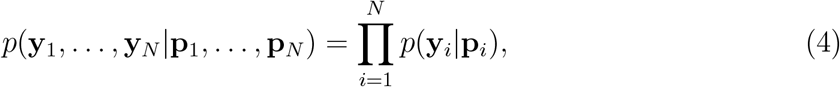

where

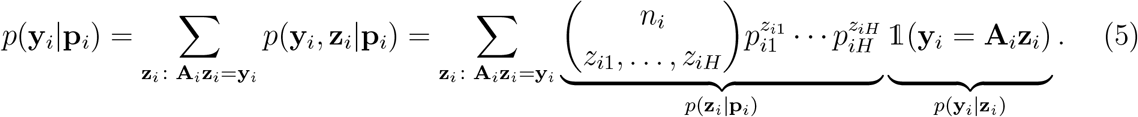

The summation in (5) requires us to enumerate, for each *i* = 1, …, *N*, the *feasible set* of solutions

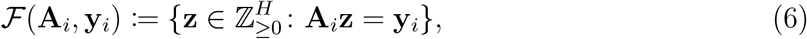

where 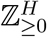is the space of *H*-dimensional nonnegative integer vectors. We find all elements of the feasible set with a branch-and-bound algorithm (Supplementary Material 1). This algorithm is run once prior to parameter inference.

### 2.1 Gaussian process prior specification

To account for the correlation between haplotype prevalences for different studies, we model the haplotype prevalences as a softmax transformation of *H* independent Gaussian processes (GPs):

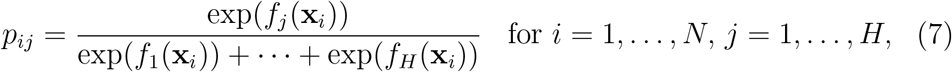

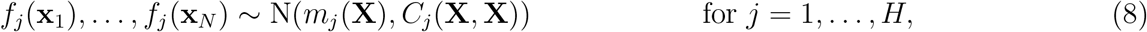

where **p**_*i*_ are the haplotype prevalences of population *i*, 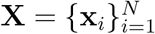are the spatiotemporal coordinates and covariates for each study, and *f*_*j*_ is the *j*th GP whose mean function and covariance function are *m*_*j*_ and *C*_*j*_ respectively. The vector *m*_*j*_(**X**) denotes the concatenation of (*m*_*j*_(**x**_1_), …, *m*_*j*_(**x**_*N*_)), and *C*_*j*_(**X, X**) is a matrix whose (*i, i*′)th entry is *C*_*j*_(**x**_*i*_, **x**_*i*′_). The mean and covariance functions are further parametrised by GP hyperparameters ***θ*** (Figure 1).

For each data point *i* = 1, …, *N*, our covariates **x**_*i*_ = (*λ*_*i*_, *ϕ*_*i*_, *t*_*i*_, *r*_*i*_) consist of the longitude *λ*_*i*_, latitude *ϕ*_*i*_, median year of study *t*_*i*_, and *P. falciparum* parasite rate *r*_*i*_ (as estimated from the Malaria Atlas Project [13]). We assume that the mean function varies linearly with the parasite rate:

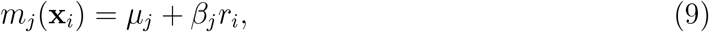

where *μ*_*j*_ is a baseline mean value and *β*_*j*_ quantifies the effect of parasite rate on the prevalence of haplotype *j*. We choose our covariance function from the Gneiting class of covariance functions on a sphere [14], along with a white noise term:

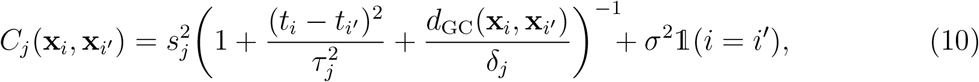

where 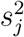 is the spatiotemporal variance, *τ*_*j*_ is the timescale parameter, *δ*_*j*_ is the lengthscale parameter, *σ*^2^ is the noise variance, *d*_GC_(**x**_*i*_, **x**_*i*′_) is the great circle distance (in degrees) between data points *i* and *i*′, and 𝟙(*·*) is the indicator function. Other Gneiting covariance functions are possible, but we choose a simple one that resembles the rational quadratic covariance function. Note that all hyperparameters except for *σ*^2^ are haplotype-specific. We place weakly informative priors on the GP hyperparameters 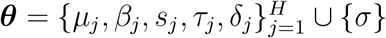; see Supplementary Material 2 for details.

### 2.1 Bayesian inference

We perform Bayesian inference using Markov chain Monte Carlo (MCMC) to obtain the posterior distribution

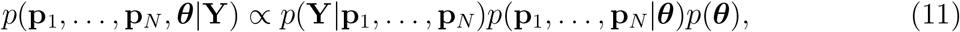

where **Y** denotes the collection of all observed data (**y**_1_, …, **y**_*N*_), the likelihood *p*(**Y**|**p**_1_, …, **p**_*N*_) is defined in (4)–(5), the prior *p*(**p**_1_, …, **p**_*N*_ |***θ***) in Section 2.2 and the hyperprior *p*(***θ***) in Supplementary Material 2. We use the No-U-Turn sampler (NUTS) [15] to perform MCMC sampling, which utilises gradient information of the logarithm of (11) to produce posterior chains of low autocorrelation. We run 5 MCMC chains each with 1000 iterations each, discarding the first half as burn-in iterations. After MCMC sampling, we produce predictive maps of haplotype prevalences on grid cells of size 0.2° × 0.2°across sub-Saharan Africa for each year between 2000 and 2020. Let **p**\* denote the vector of haplotype prevalences for an arbitrary grid cell with covariates **x**\*. To obtain samples from the posterior *p*(**p**\*|**Y**), we draw samples from the distribution *p*(**p**\*|**p**_1_, …, **p**_*N*_, ***θ***), where the values of **p**_1_, …, **p**_*N*_, ***θ*** are taken from the posterior samples output by NUTS. This is justified by the fact that

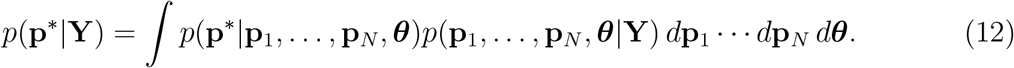

Based on GP theory [16], the distribution *p*(**p**\*|**p**_1_, …, **p**_*N*_, ***θ***) follows a normal distribution with the softmax transformation (7) applied, allowing the posterior predictive distribution *p*(**p**\*|**Y**) to be exactly sampled given samples of the posterior distribution *p*(**p**_1_, …, **p**_*N*_, ***θ***|**Y**).

## 3 Results

We fit our latent multinomial GP model to SP resistance data in Africa, where it is common for parasites to have multiple SP-resistant mutations on the *dhps* gene and studies may report the number of samples with mutations differently. Here we consider *G* = 3 mutations on the *dhps* gene, namely A437G, K540E, and A581G, leading to *H* = 8 distinct full haplotypes. We use a dataset collated by the Worldwide Antimalarial Resistance Network [12], as detailed in Supplementary Material 3. Our primary goal is to obtain Bayesian estimates of the prevalences (i.e. multinomial probabilities) of the 8 full haplotypes across sub-Saharan Africa over the duration of interest 2000–2020.

The total computational time for preprocessing and MCMC was 10.5 hours. We achieve a potential scale reduction factor [17] of 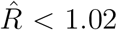 for all parameters, and each haplotype prevalence had an effective sample size greater than 500. The MCMC chains exhibit good mixing, as indicated by the representative trace plots shown in Supplementary Material 4.

We divide the region of interest into a 0.2°× 0.2°grid, where we define our area of interest to be the region where the Malaria Atlas Project maps malaria transmission [13, 18]. Foreach spatiotemporal coordinate **x**\* of a grid cell and year, we find the posterior predictive distribution of the full haplotype prevalences at **x**\*. Figure 2 shows the distribution of posterior median prevalences over the region of interest for all 8 haplotypes during 2000–2020.

**Figure 2.**
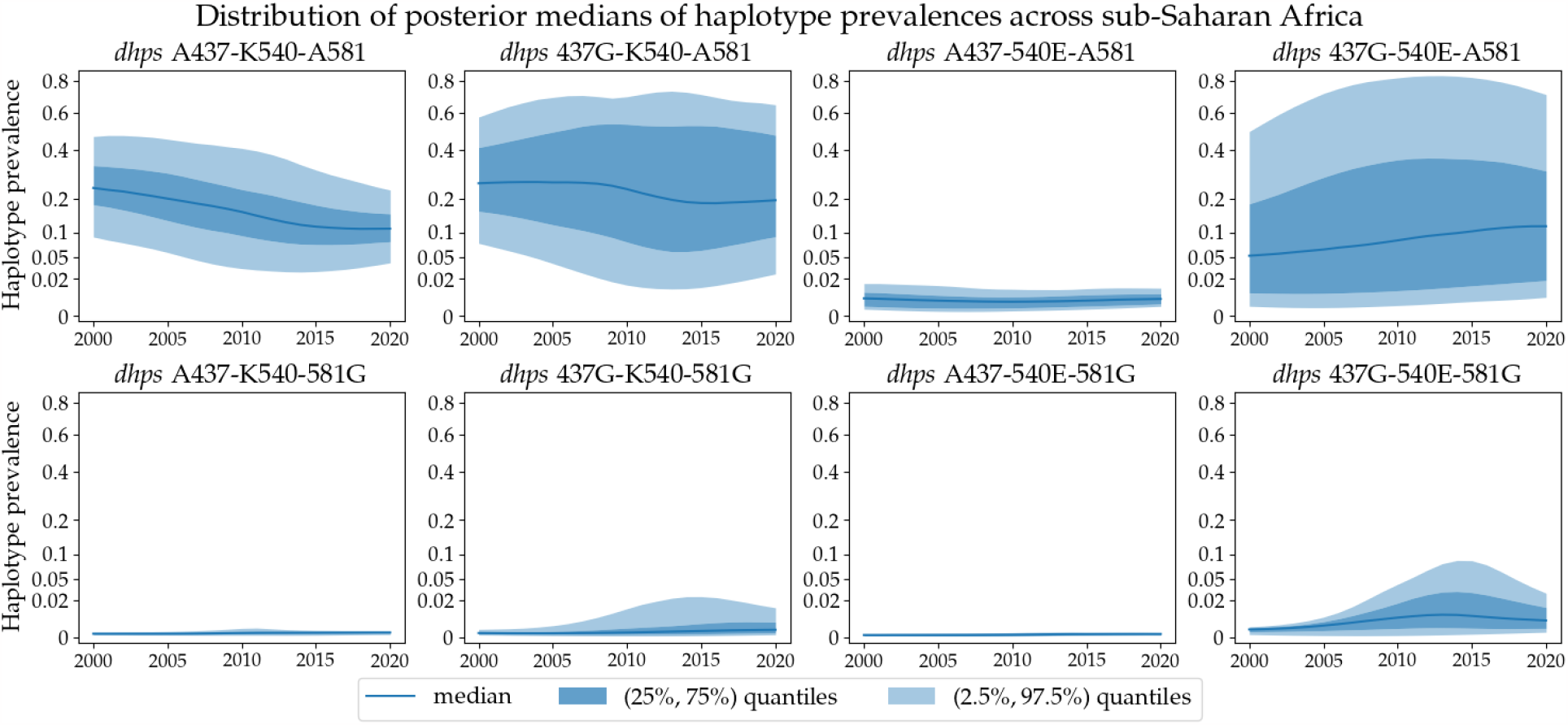
Distribution (over grid cells of study area) of the posterior medians of *H* = 8 prevalences of *dhps* haplotypes from year 2000 to year 2020. The dark blue line represents the median over the grid cells of our study area, the dark blue band represents the 50% interval and the light blue band the 95% interval. Posterior medians in all figures are presented on a square root scale.

Over this region, of the *H* = 8 haplotypes, three have very low prevalence over the entire duration of interest; A437-540E-A581, A437-K540-581G and A437-540E-581G.

We also provide visual summaries of the spatial distribution of prevalences of 5 selected haplotypes (the three low prevalence haplotypes from Figure 2 are excluded) and their change over time. Figure 3 shows the median and standard deviation summaries of the posterior predictive distributions over the region of interest in the years 2000, 2010, and 2020. The results presented in this figure are broadly consistent with literature in that the vast majority of mutant *dhps* haplotypes have the A437G mutation [19]. The spatial patterns shown by our heatmaps corroborate with the results of Naidoo and Roper [20], who report the double mutation A437G-K540E as the most prevalent mutant haplotype in East Africa (third column), and also the single mutation A437G as the most prevalent mutant haplotype in West and Central Africa (second column).

**Figure 3.**
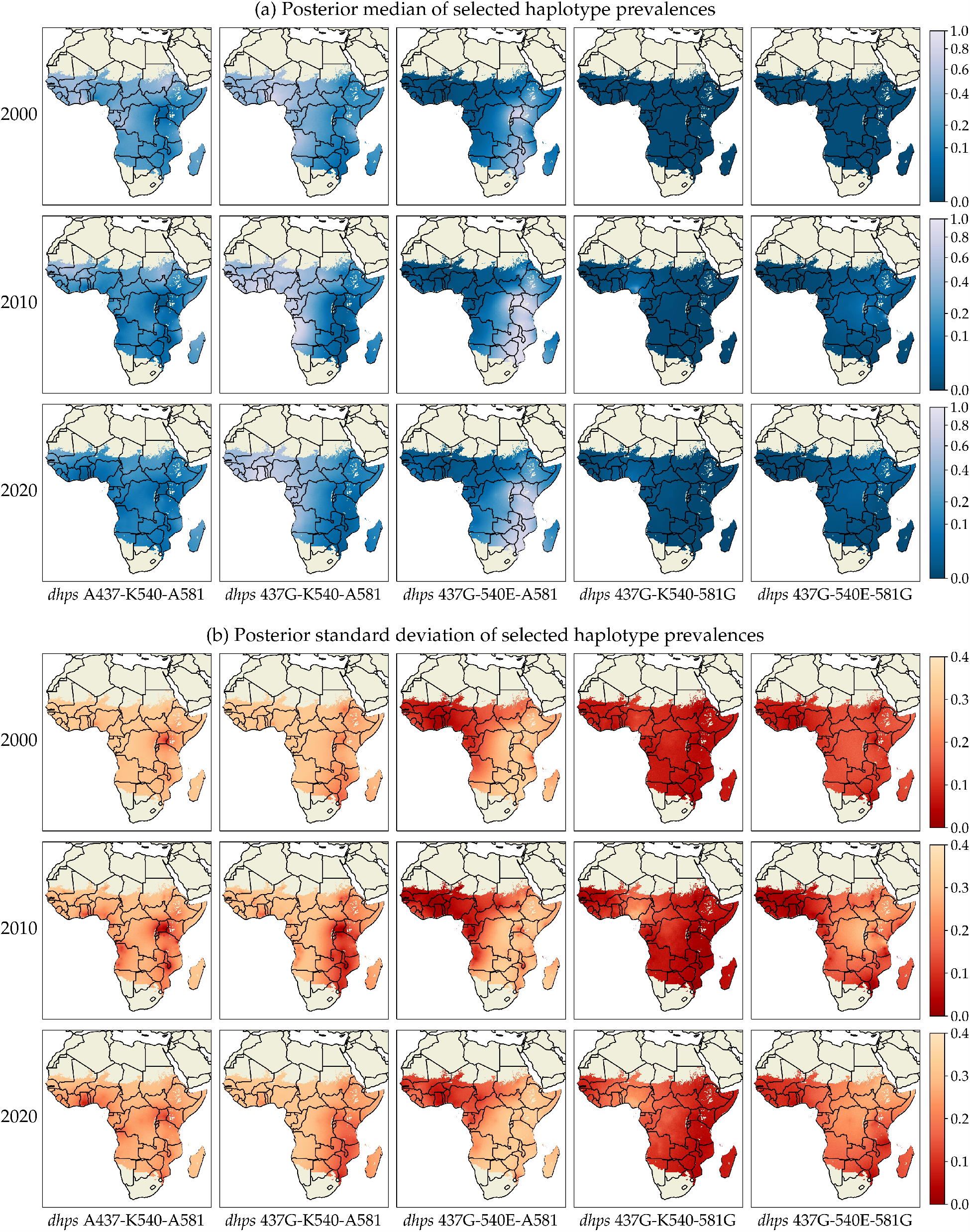
(a) Posterior median and (b) posterior standard deviation for prevalences of selected *dhps* haplotypes in years 2000 (top row), 2010 (middle row) and 2020 (bottom row).

In the case where the prevalence of an individual mutation (A437G, K540E, or A581G) is of interest, these can still be captured as outputs of our model by simply summing over the halplotypes that include the mutation. In Figure 4, the spatial distribution of A437G, K540E, and A581G are summarised with posterior median (top row) and standard deviation (bottom row) in 2020. These results are broadly consistent with results shown in Flegg et al. [6] for the same three markers in 2020.

**Figure 4.**
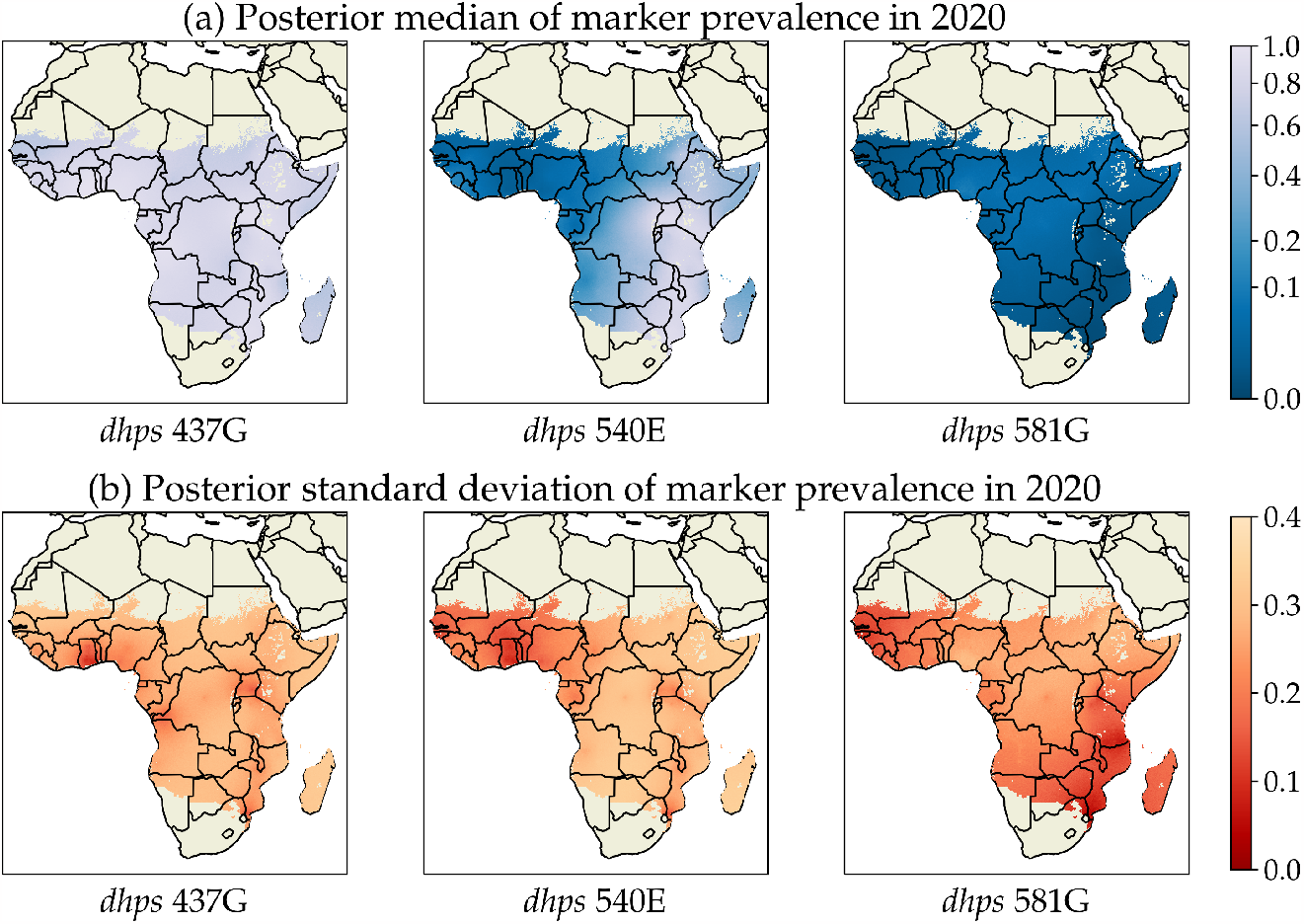
Posterior summary for prevalences of selected *dhps* individual markers in year 2020. The top row shows the posterior median for *dhps* 437, 540, 581; the bottom row shows the posterior standard deviation for *dhps* 437, 540, 581.

## 4 Discussion

In this paper, we develop a spatiotemporal geostatistical model to infer for the first time the prevalence of multi-marker drug-resistant malaria. We illustrate the utility of this new model for SP, which is a commonly used drug for intermittent preventive treatment of malaria in pregnancy, children and infants. Since drug-resistant haplotypes and markers are often used as a proxy for treatment efficacy, these maps can help inform antimalarial drug policies. Our methods take on a Bayesian approach, which are able to quantify uncertainties about the prevalence of the drug resistance haplotypes. A benefit of quantifying uncertainty is its use in optimising sampling strategies for future monitoring of drug resistance [21]. Existing geostatistical methods in spatiotemporal modelling of antimalarial drug resistance use a binomial likelihood [3, 6, 21, 22], which can only infer the prevalence of a single category e.g. prevalence of one mutation, or prevalence of the wildtype haplotype. Our models are capable of handling multiple haplotypes simultaneously by using a multinomial likelihood, leading to more refined inference about drug resistance. This has not been done in previous work, as not all studies provide data on full haplotypes; studies may only report on a subset of mutations, or group haplotypes by the number of mutations present, or provide counts for each mutation separately. We were able to handle these types of partially-reported data by using a latent multinomial model that treats each reported category as a subset of all full haplotypes. Although the counts of each full haplotype are not all experimentally determined, our approach of enumerating all possible latent counts allows us to leverage the partially-observed data for inferring the prevalences of the full haplotypes.

One limitation of our work presented here is that sampling bias may be present due to population heterogeneity. For example, since SP is commonly used for intermittent preventive treatment of malaria, many studies in SP resistance take blood samples from pregnant people. Bias may also arise from the choice of mutations reported. If the prevalence of a mutation is very low, it is less likely to be reported by a study. This may lead to an overestimation in the prevalence of mutations that are not often reported. Another limitation is a lack of model checking to verify whether our model fits the data adequately. Since the haplotype categories reported are inconsistent across studies, it is not straightforward what model checking procedures should be applied.

A possible extension to our approach is to include more covariates, such as demographic or environmental covariates, e.g. human mobility data. However, using more covariates implies that more model parameters need to be inferred. Our current MCMC approach is already computationally expensive, an issue that may be exacerbated by the inclusion of more covariates. For a dataset with more markers and/or larger pools, our enumeration approach may become infeasible, as there are too many possible latent count solutions to enumerate. In this case, we can instead treat the latent counts as model parameters to be sampled during MCMC using a custom MCMC sampler [10] based on Markov bases [23]. MCMC sampling of the latent counts cannot be performed by gradient-based samplers such as NUTS, as they cannot handle discrete model parameters. There is also more work to do in extending the model to consider dependent GPs, as it is possible that the prevalences of different haplotypes are related to each other, using for example the linear model of coregionalisation [24] or the semiparametric latent factor model [25].

At present, the World Health Organisation provides recommendations for implementing intermittent preventive treatment in pregnancy with SP based on the prevalence of the *dhps* K540E and A581G mutations [26]. Although it is known that different *dhps* haplotypes confer different degrees of SP resistance [20], these recommendations are based on the prevalence of individual mutations, rather than that of full haplotypes. One possible reason is that there are no existing methods in literature to infer the prevalence of full haplotypes from partially-reported data. We address this gap in literature by describing how a latent multinomial GP model can be used to produce spatiotemporal maps of these prevalences. The results we present in this paper are able to quantify the spread of various drug-resistant haplotypes, and provide uncertainty estimates that can help optimise sampling strategies for future monitoring of antimalarial drug resistance.

## Data availability

The dataset supporting this article is available at www.wwarn.org/tracking-resistance/sp-molecular-surveyor. The code used for analysis is available at github.com/ysfoo/haplm.

## References

[1] Sibley CH, Hyde JE, Sims PF, Plowe CV, Kublin JG, Mberu EK, Cowman AF, Winstanley PA, Watkins WM, and Nzila AM. Pyrimethamine–sulfadoxine resistance in Plasmodium falciparum: what next? Trends in Parasitology. 2001. 17 (12): pp. 582–588.

[2] Nsanzabana C, Djalle D, Güerin PJ, Müenard D, and Gonzälez IJ. Tools for surveillance of anti-malarial drug resistance: an assessment of the current landscape. Malaria journal. 2018. 17: pp. 1–16.

[3] Flegg JA, Patil AP, Venkatesan M, Roper C, Naidoo I, Hay SI, Sibley CH, and Guerin PJ. Spatiotemporal mathematical modelling of mutations of the dhps gene in African Plasmodium falciparum. Malaria Journal. 2013. 12 (1): p. 249.

[4] Deutsch-Feldman M et al. The changing landscape of Plasmodium falciparum drug resistance in the Democratic Republic of Congo. BMC Infectious Diseases. 2019. 19 (1): p. 872.

[5] Amimo F, Lambert B, Magit A, Sacarlal J, Hashizume M, and Shibuya K. Plasmodium falciparum resistance to sulfadoxine-pyrimethamine in Africa: a systematic analysis of national trends. BMJ Global Health. 2020. 5 (11): e003217.

[6] Flegg JA, Humphreys GS, Montanez B, Strickland T, Jacome-Meza ZJ, Barnes KI, Raman J, Guerin PJ, Hopkins Sibley C, and Dahlström Otienoburu S. Spatiotemporal spread of Plasmodium falciparum mutations for resistance to sulfadoxine-pyrimethamine across Africa, 1990–2020. PLOS Computational Biology. 2022. 18 (8): e1010317.

[7] Gesase S, Gosling RD, Hashim R, Ord R, Naidoo I, Madebe R, Mosha JF, Joho A, Mandia V, Mrema H, et al. High resistance of Plasmodium falciparum to sulphadoxine/pyrimethamine in northern Tanzania and the emergence of dhps resistance mutation at Codon 581. PLoS One. 2009. 4 (2): e4569.

[8] Harrington W, Mutabingwa T, Muehlenbachs A, Sorensen B, Bolla M, Fried M, and Duffy P. Competitive facilitation of drug-resistant Plasmodium falciparum malaria parasites in pregnant women who receive preventive treatment. Proceedings of the National Academy of Sciences. 2009. 106 (22): pp. 9027–9032.

[9] Link WA, Yoshizaki J, Bailey LL, and Pollock KH. Uncovering a Laten Multinomial: Analysis of Mark-Recapture Data with Misidentification. Biometrics. 2010. 66 (1): pp. 178–185.

[10] Foo YS and Flegg JA. Haplotype frequency inference from pooled genetic data with a latent multinomial model. 2023. arXiv: 2308.16465 [stat.ME].

[11] Triglia T, Wang P, Sims PF, Hyde JE, and Cowman AF. Allelic exchange at the endogenous genomic locus in Plasmodium falciparum proves the role of dihydropteroate synthase in sulfadoxine-resistant malaria. The EMBO journal. 1998. 17 (14): pp. 3807–3815.

[12] Infectious Diseases Data Observatory. SP Molecular Surveyor. 2022. URL: https://www.wwarn.org/tracking-resistance/sp-molecular-surveyor (visited on 06/18/2023).

[13] Malaria Atlas Project. Data. URL: https://malariaatlas.org/ (visited on 05/20/2023).

[14] Porcu E, Furrer R, and Nychka D. 30 years of space–time covariance functions. WIREs Computational Statistics. 2021. 13 (2): e1512.

[15] Hoffman MD and Gelman A. The No-U-Turn Sampler: Adaptively Setting Path Lengths in Hamiltonian Monte Carlo. Journal of Machine Learning Research. 2014. 15 (47): pp. 1593–1623.

[16] Rasmussen CE and Williams CKI. Gaussian Processes for Machine Learning (Adaptive Computation and Machine Learning). The MIT Press, 2005. ISBN: 026218253X.

[17] Vehtari A, Gelman A, Simpson D, Carpenter B, and Bürkner PC. Rank-Normalization, Folding, and Localization: An Improved R^ for Assessing Convergence of MCMC (with Discussion). Bayesian Analysis. 2021. 16 (2): pp. 667–718.

[18] Weiss DJ, Lucas TC, Nguyen M, Nandi AK, Bisanzio D, Battle KE, Cameron E, Twohig KA, Pfeffer DA, Rozier JA, et al. Mapping the global prevalence, incidence, and mortality of Plasmodium falciparum, 2000–17: a spatial and temporal modelling study. The Lancet. 2019. 394 (10195): pp. 322–331.

[19] Vinayak S et al. Origin and Evolution of Sulfadoxine Resistant Plasmodium falciparum. PLoS Pathogens. 2010. 6 (3): e1000830.

[20] Naidoo I and Roper C. Mapping ‘partially resistant’, ‘fully resistant’, and ‘super resistant’ malaria. Trends in Parasitology. 2013. 29 (10): pp. 505–515.

[21] Grist EPM et al. Optimal health and disease management using spatial uncertainty: a geographic characterization of emergent artemisinin-resistant Plasmodium falciparum distributions in Southeast Asia. International Journal of Health Geographics. 2016. 15 (1): p. 37.

[22] Tun KM et al. Spread of artemisinin-resistant Plasmodium falciparum in Myanmar: a cross-sectional survey of the K13 molecular marker. The Lancet Infectious Diseases. 2015. 15 (4): pp. 415–421.

[23] Schofield MR and Bonner SJ. Connecting the latent multinomial. Biometrics. 2015. 71 (4): pp. 1070–1080.

[24] Journel AG and Huijbregts CJ. Mining geostatistics. English. Academic Press, London, 1978.

[25] Teh YW, Seeger M, and Jordan MI. Semiparametric latent factor models. In: Cowell RG and Ghahramani Z, eds. Proceedings of the Tenth International Workshop on Artificial Intelligence and Statistics; Jan. 6–8, 2005; Barbados. Oxford, UK: PMLR; 2005. Pp. 333–340.

[26] World Health Organisation. Meeting report of the Evidence Review Group on intermittent preventive treatment (IPT) of malaria in pregnancy. Geneva, Switzerland, 2013.

