## Supplementary Material (text) for "A spatiotemporal model of multi-marker antimalarial resistance"

#### 1 Algorithm for finding the feasible set

In this section, we describe a branch-and-bound algorithm for solving $\mathbf{A}\mathbf{z} = \mathbf{y}$ over nonnegative integers $z_1, \ldots, z_H$ given a binary matrix $\mathbf{A} \in \{0, 1\}^{R \times H}$ and nonnegative integers $y_1, \ldots, y_R$. Note that the index $i$ from the main text is dropped for conciseness here. We assume that the condition $z_1 + \cdots + z_H = n$ is encoded in the linear system $\mathbf{A}\mathbf{z} = \mathbf{y}$ (see Section 2.1 of main text). If the configuration matrix $\mathbf{A}$ is not of full row rank, we use row reduction to obtain a submatrix consisting of a maximal set of linearly independent rows. This removes redundant information observed from a latent multinomial model, see Zhang et al. (2019) for an explanation of why inference results are not affected by this procedure. Thus, assuming that the configuration matrix $\mathbf{A}$ is of full row rank, we can find $R$ columns of $\mathbf{A}$ that are linearly independent. Without loss of generality, we rearrange the columns of $\mathbf{A}$ such that these $R$ linearly independent columns are the last $R$ columns, denoted as $\mathbf{A}_{H-R+1:H}$. Since $\mathbf{y} = \mathbf{A}_{1:H-R}\mathbf{z}_{1:H-R} + \mathbf{A}_{H-R+1:H}\mathbf{z}_{H-R+1:H}$, it follows that

$$\mathbf{z}_{H-R+1:H} = \mathbf{A}_{H-R+1:H}^{-1}(\mathbf{y} - \mathbf{A}_{1:H-R}\mathbf{z}_{1:H-R}), \tag{1}$$

where $\mathbf{A}_{1:H-R}, \mathbf{z}_{1:H-R}, \mathbf{z}_{H-R+1:H}$ denotes the first $H-R$ columns of $\mathbf{A}$, the first $H-R$ entries of $\mathbf{z}$, the last $R$ entries of $\mathbf{z}$ respectively. To find all solutions to the system, we perform a branch-and-bound search to find all possible values of $z_1, \ldots, z_{H-R}$. Starting from $h = 1$, the algorithm branches on an interval of possible values for $z_h$ and increments $h$ whenever a branch is travelled down. If this succeeds until $h = H - R$, we then find the last $R$ entries of $\mathbf{z}$ by using (1). If the result consists of nonnegative integers, we accept $\mathbf{z}$ as a solution to $\mathbf{A}\mathbf{z} = \mathbf{y}$. We then backtrack the search path (decrementing $h$), and explore all other branches to find other solutions. The search is made efficient by finding lower and upper bounds for $z_h$ based on the values of $z_1, \ldots, z_{h-1}$ when branching on the value of $z_h$ for $h = 1, \ldots, H$.

Before the search procedure, we first determine preliminary lower bounds $l_h$ and upper bounds $u_h$ for each entry $z_h$ that are satisfied by all nonnegative integer solutions to $\mathbf{A}\mathbf{z} = \mathbf{y}$. A simple choice is to set

$$l_h = 0, \qquad u_h = \min_{r=1,\ldots,R}\{a_{r,h}y_r + (1 - a_{r,h})(n - y_r)\}. \tag{2}$$

The lower bound is trivial, whereas the upper bound is true because the $r$th equation in the system implies that $z_h \leq y_r$ if $a_{r,h} = 1$, or $z_h \leq n - y_r$ if $a_{r,h} = 0$. For each $h = 1, \ldots, H$, we now seek to derive bounds for $z_h$ using the values of $z_1, \ldots, z_{h-1}$ along the current search path. For any fixed $r$ and $h$, we have

$$
\begin{aligned}
z_h &= l_h + (z_h - l_h) \\
&\leq l_h + \sum_{h'=h}^{H} \mathbb{1}(a_{r,h'} = a_{r,h})(z_{h'} - l_{h'}) \\
&= \begin{cases} y_r - \sum_{h'=1}^{h-1} a_{r,h'} z_{h'} - \sum_{h'=h+1}^{H} a_{r,h'} l_{h'} & \text{if } a_{r,h} = 1, \\ n - y_r - \sum_{h'=1}^{h-1} (1 - a_{r,h'}) z_{h'} - \sum_{h'=h+1}^{H} (1 - a_{r,h'}) l_{h'} & \text{if } a_{r,h} = 0. \end{cases}
\end{aligned} \tag{3}
$$

The inequality holds since $z_{h'} \geq l_{h'}$, while the last equality holds because of $y_r = \sum_{h'=1}^{H} a_{r,h'} z_{h'}$ and $n - y_r = \sum_{h'=1}^{H} (1 - a_{r,h'}) z_{h'}$. We define

$$
\begin{aligned}
U_1(r; h, z_1, \ldots z_{h-1}) &= y_r - \sum_{h'=1}^{h-1} a_{r,h'} z_{h'} - \sum_{h'=h+1}^{H} a_{r,h'} l_{h'}, \\
U_0(r; h, z_1, \ldots z_{h-1}) &= n - y_r - \sum_{h'=1}^{h-1} (1 - a_{r,h'}) z_{h'} - \sum_{h'=h+1}^{H} (1 - a_{r,h'}) l_{h'},
\end{aligned}
$$

to write the inequality in (3) more concisely as $z_h \leq U_{a_{r,h}}(r; h, z_1, \ldots z_{h-1})$. We similarly define

$$
\begin{aligned}
L_1(r; h, z_1, \ldots z_{h-1}) &= y_r - \sum_{h'=1}^{h-1} a_{r,h'} z_{h'} - \sum_{h'=h+1}^{H} a_{r,h'} u_{h'}, \\
L_0(r; h, z_1, \ldots z_{h-1}) &= n - y_r - \sum_{h'=1}^{h-1} (1 - a_{r,h'}) z_{h'} - \sum_{h'=h+1}^{H} (1 - a_{r,h'}) u_{h'},
\end{aligned}
$$

to obtain the inequality $z_h \geq L_{a_{r,h}}(r; h, z_1, \ldots z_{h-1})$.

The branch-and-bound algorithm is given in Algorithm S1. The values for $U_1, U_0, L_1, L_0$ are initialised in lines 4–8, where $h = 1$ and $r = 1, \ldots, R$. Given the values of $z_1, \ldots, z_{h-1}$ on the current search path, the algorithms finds lower and upper bounds for $z_h$ in lines 11–12 using the inequality $L_{a_{r,h}}(r; h, z_1, \ldots z_{h-1}) \leq z_h \leq U_{a_{r,h}}(r; h, z_1, \ldots z_{h-1})$ over $r = 1, \ldots, R$. The branching occurs in lines 19–24, where $U_1, U_0, L_1, L_0$ are updated based on the chosen value of $z_h$.

If the actual range of values that $z_h$ can take is much narrower than the interval $[l_h, u_h]$ as defined in (2), it may be computationally more efficient to find the actual minimum and maximum values that $z_h$ can take, i.e. setting

$$
\begin{aligned}
l_h &= \min\{z_h : \mathbf{Az} = \mathbf{y}, z_1 \geq 0, \ldots, z_H \geq 0\}, \\
u_h &= \max\{z_h : \mathbf{Az} = \mathbf{y}, z_1 \geq 0, \ldots, z_H \geq 0\}.
\end{aligned} \tag{4}
$$

for each $h = 1, \dots, H$. These optimisation problems can be solved using integer linear programming. This introduces a computational overhead before the branch-and-bound search, but prunes the search space as $\mathbf{z}$ would have tighter bounds.

**Algorithm S1:** Branch-and-bound search for integer linear system with 0-1 coefficients over nonnegative integers with known sum

**Input:** $\mathbf{y}, \mathbf{A}, n, l_1, \dots, l_H, u_1, \dots, u_H$
**Output:** $\mathcal{S}$, a set of nonnegative integer solutions $\mathbf{z}$ to $\mathbf{Az} = \mathbf{y}$

$\mathcal{S} \leftarrow \{\}$
$\mathbf{z} \leftarrow$ empty vector of size $H$
$U_1, U_0, L_1, L_0 \leftarrow$ empty $R \times (H - R)$ array
**for** $r \leftarrow 1$ **to** $R$ **do**
  $U_1[r, 1] \leftarrow y_r - \sum_{h=2}^{H} a_{r,h} l_h$
  $U_0[r, 1] \leftarrow n - y_r - \sum_{h=2}^{H} (1 - a_{r,h}) l_h$
  $L_1[r, 1] \leftarrow y_r - \sum_{h=2}^{H} a_{r,h} u_h$
  $L_0[r, 1] \leftarrow n - y_r - \sum_{h=2}^{H} (1 - a_{r,h}) u_h$
compute $\mathbf{A}_{H-R+1:H}^{-1}$
**Function** `branch(`$h$`)`**:**
  $z_{\min} = \max\big(l_h, \max_{r=1,\dots,R} L_{a_{r,h}}[r, h]\big)$
  $z_{\max} = \min\big(u_h, \min_{r=1,\dots,R} U_{a_{r,h}}[r, h]\big)$
  **if** $h = H - R$ **then**
    **for** $z_h \leftarrow z_{min}$ **to** $z_{max}$ **do**
      $\mathbf{z}_{H-R+1:H} \leftarrow \mathbf{A}_{H-R+1:H}^{-1}(\mathbf{y} - \mathbf{A}_{1:H-R}\mathbf{z}_{1:H-R})$
      **if** *all entries of* $\mathbf{z}_{H-R+1:H}$ *are nonnegative integers* **then**
        $\mathcal{S} \leftarrow \mathcal{S} \cup \{\mathbf{z}\}$
  **else**
    **for** $z_h \leftarrow z_{min}$ **to** $z_{max}$ **do**
      **for** $r \leftarrow 1$ **to** $R$ **do**
        $U_1[r, h+1] \leftarrow U_1[r, h] - a_{r,h} z_h + a_{r,h+1} l_{h+1}$
        $U_0[r, h+1] \leftarrow U_0[r, h] - (1 - a_{r,h}) z_h + (1 - a_{r,h+1}) l_{h+1}$
        $L_1[r, h+1] \leftarrow L_1[r, h] - a_{r,h} z_h + a_{r,h+1} u_{h+1}$
        $L_0[r, h+1] \leftarrow L_0[r, h] - (1 - a_{r,h}) z_h + (1 - a_{r,h+1}) u_{h+1}$
      `branch(`$h + 1$`)`
  **return**
`branch(1)`
**return** $\mathcal{S}$

### 2 Priors for Gaussian process hyperparameters

In Section 2.2 of the main text, we specified the mean function and covariance function for each haplotype $j = 1, \dots, H$ to be

$$m_j(\mathbf{x}_i) = \mu_j + \beta_j r_i, \tag{5}$$

and

$$C_j(\mathbf{x}_i, \mathbf{x}_{i'}) = s_j^2\left(1 + \frac{(t_i - t_{i'})^2}{\tau_j^2} + \frac{d_{\mathrm{GC}}(\mathbf{x}_i, \mathbf{x}_{i'})}{\delta_j}\right)^{-1} + \sigma^2 \mathbb{1}(i = i'), \tag{6}$$

respectively, where $r_i$ is the *P. falciparum* parasite rate at $\mathbf{x}_i$, $\mu_j$ is a baseline mean value, $\beta_j$ quantifies the effect of parasite rate on the prevalence of haplotype $j$, $s_j^2$ is the spatiotemporal variance, $\tau_j$ is the timescale parameter, $\delta_j$ is the lengthscale parameter, $\sigma^2$ is the noise variance, and $\mathbb{1}(\cdot)$ is the indicator function.

In this section, we specify priors for the Gaussian process hyperparameters

$$\boldsymbol{\theta} = \{\mu_j, \beta_j, s_j, \tau_j, \delta_j\}_{j=1}^H \cup \{\sigma\}.$$

Note that if all entries of $\boldsymbol{\mu} = \{\mu_j\}_{j=1}^H$ across $j = 1, \ldots, H$ are incremented by the same value, this keeps the distribution of $(\mathbf{p}_1, \ldots, \mathbf{p}_N)$ unchanged due to the softmax transformation (Equation 7 of the main text). To improve identifiability of $\mu_j$, we impose a sum-to-zero constraint $\mu_1 + \cdots + \mu_H = 0$. We also apply the same sum-to-zero constraint for $\mathbf{r} = \{r_j\}_{j=1}^H$ for similar reasons. We satisfy these constraints by conditioning a random vector following the normal distribution $\mathrm{N}(\mathbf{0}_H, 2^2\mathbf{I}_H)$ to have its entries sum to zero (Fraser, 1951), resulting in the prior distributions

$$\boldsymbol{\mu} \sim \mathrm{N}\left(\mathbf{0}_H, 2^2\left(\mathbf{I}_H - \frac{1}{H}\mathbf{J}_H\right)\right), \tag{7}$$

$$\mathbf{r} \sim \mathrm{N}\left(\mathbf{0}_H, 2^2\left(\mathbf{I}_H - \frac{1}{H}\mathbf{J}_H\right)\right), \tag{8}$$

where $\mathbf{0}_H$ is a vector of $H$ zeros, $\mathbf{I}_H$ is the $H \times H$ identity matrix, and $\mathbf{J}_H$ is a $H \times H$ matrix of ones.

As for the covariance function hyperparameters, we place weakly informative inverse gamma priors (shape-scale parametrisation)

$$s_1, \ldots, s_H \sim \mathrm{IG}(3, 3) \tag{9}$$

$$\tau_1, \ldots, \tau_H \sim \mathrm{IG}(3, 5) \tag{10}$$

$$\delta_1, \ldots, \delta_H \sim \mathrm{IG}(3, 7) \tag{11}$$

$$\sigma \sim \mathrm{IG}(3, 1). \tag{12}$$

We choose inverse gamma priors as they suppress values around zero and infinity, and the choice of the shape and scale parameters are informed by the probable values of each hyperparameter. Specifically, the following events (i.e. range of probable values for each hyperparameter) each have a 0.99 prior probability of occurring:

$$0.32 < s_j < 8.85$$
$$0.54 < \tau_j < 14.52 \text{ (years)}$$
$$0.76 < \delta_j < 21.16 \text{ (degrees)}$$
$$0.11 < \sigma < 2.90.$$

### 3 Data inclusion

We retrieved from www.wwarn.org/tracking-resistance/sp-molecular-surveyor prevalence data for *dhps* 437, 540, 581 in sub-Saharan Africa since 2000. We treat mixed infections as mutations. We remove data points where the study site is absent, or the study start and end years differ by at least 4 years. Such points are considered to not have sufficient spatial or temporal resolution, so we deem them to be not reliable enough for inference. This results in 256 data points.

For each data point, we then attempt to find the feasible set given the observed counts, i.e. enumerate all possible latent counts. We find that the feasible set is empty for 15 data points, which we subsequently remove — an empty feasible set indicates that the observed counts are erroneous as there is no latent count vector that could have produced such observed counts. Such errors can arise if the number of samples successfully haplotyped for each set of mutations are different, but there is no clear indication of such discrepancies in the dataset description. Our final dataset used for inference consists of $N = 241$ data points.

### 4 Supplementary figures

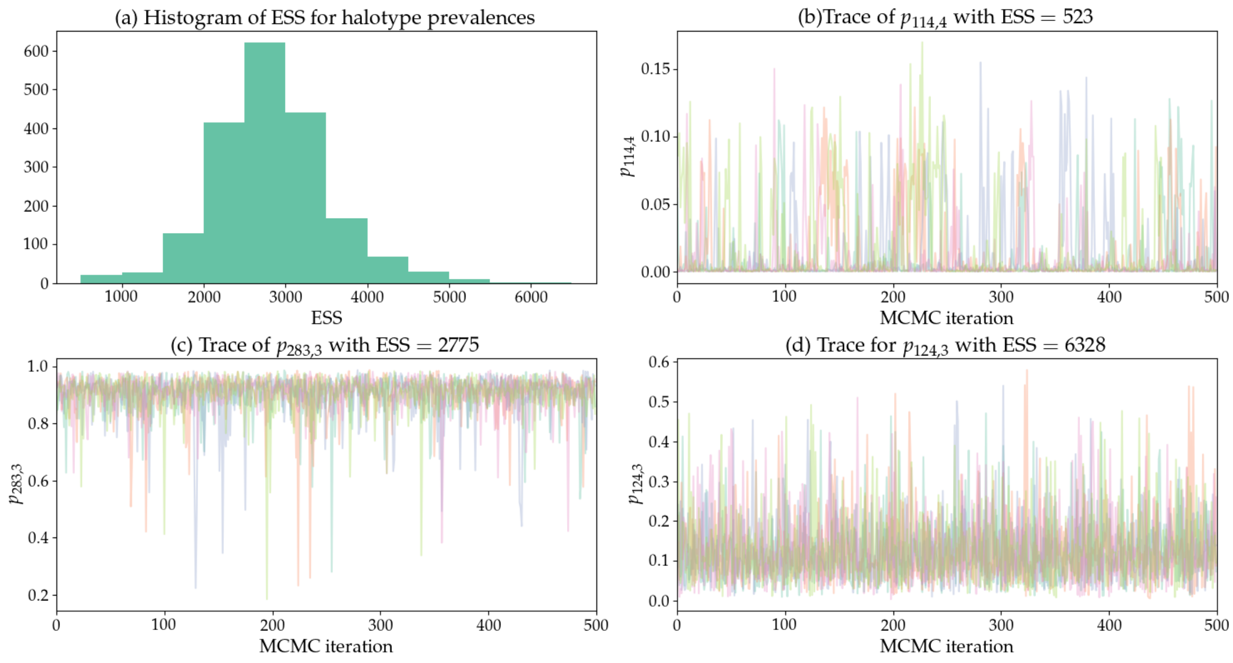


Figure S1: (a) Histogram of effective sample size (ESS) values for haplotype prevalences $p_{ij}$ over all data points $i = 1, \ldots, N$ and haplotypes $j = 1, \ldots, H$. The trace plots depict representative MCMC traces for the haplotype prevalence $p_{ij}$ with the (b) minimum ESS, (c) median ESS, (d) maximum ESS.

### References

Fraser, D. A. S. (1951). Normal samples with linear constraints and given variances. *Canadian Journal of Mathematics*, *3*, 363–366.

Zhang, W., Bravington, M. V., & Fewster, R. M. (2019). Fast likelihood-based inference for latent count models using the saddlepoint approximation. *Biometrics*, *75*(3), 723–733.
